# Patient consent for medical student pelvic exams under anesthesia: an exploratory retrospective chart review

**DOI:** 10.1101/2023.04.17.23288694

**Authors:** Jessica A. Jushchyshyn, Lakeisha Mulugeta-Gordon, Cara Curley, Florencia Greer Polite, Jon F. Merz

**Affiliations:** Beth Israel Deaconess Medical Center, 1309 Beacon, Office 214, Brookline, MA 02446; Department of Obstetrics & Gynecology, Hospital of the University of Pennsylvania, Philadelphia, PA 19104; Department of Obstetrics & Gynecology, Perelman School of Medicine at the University of Pennsylvania; Division of General Obstetrics & Gynecology, Perelman School of Medicine at the University of Pennsylvania, 3701 Market Street, Philadelphia, PA 19104; Medical Ethics & Health Policy, Perelman School of Medicine at the University of Pennsylvania, Blockley Hall Floor 14, 423 Guardian Drive, Philadelphia, Pennsylvania 19104-4884

## Abstract

**Background:** Legal requirements and clinical practices of securing patient consent for medical student pelvic examinations under anesthesia (EUA) vary widely, while ethical arguments and patients’ preferences for being asked for consent are well known.

**Objective:** This study was performed to examine patients’ choices to permit or refuse medical student pelvic EUAs during planned gynecologic procedures.

**Study Design:** An exploratory retrospective chart review of electronic consent forms at a single academic medical center, using contingency table and logistic regression to explore relationships between patient and provider characteristics and consent.

**Results:** Electronic consent forms were downloaded for a census of 4000 patients undergoing gynecologic surgery from September 2020 through calendar year 2022 and linked to anonymized medical record information, including patient age, race, religion, and insurance carrier, along with physician name. Physicians were coded by gender, departmental affiliation, and status (attending, resident, or fellow). Of the 4000 patients, 142 (3.6%) patients were removed from analysis because of uncertainty about the EUA consent. Of the remainder, 308 (8.0%) were asked for EUA consent more than once. Overall, of 3858 patients, 3308 (85.7%) consented every time asked and 550 (14.2%) refused or limited EUA consent at least once. Nine patients limited their consent to female students, and 2 patients refused medical student participation at all. Of the 308 asked more than once, 46 were not consistent. Exploratory multiple logistic regression Consent for pelvic exams under anesthesia analysis showed that patients identifying as Black or African American (OR=0.482, p<0.001) or Asian (OR=0.303, p<0.001), or of Moslem/Muslim/Islamic faith (OR=0.598, p=0.008) were substantially less likely to grant EUA consent than other patients. Moreover, male physicians were somewhat more likely to secure consent from patients than their female colleagues (OR=1.427, p=0.016).

**Conclusions:** The finding that some patients are more likely than others to refuse a pelvic EUA magnifies the dignitary harm from a nonconsensual invasion of intimate bodily integrity and perpetuates the historic wrongs visited upon vulnerable people of color and religious minorities. Patient’s rights to control over their own bodies can only be respected if their physicians take seriously the ethical obligation to inform their patients and ask them for permission.

## Introduction

Medical students typically perform examinations of patients for training purposes, normally securing permission from patients before doing so. It has not always been so, as ethical and legal standards for informed consent evolved only over the last 6 decades.^1-3^ One continuing exception to this regards the performance of pelvic and prostate exams, which are often conducted on unconscious, sedated patients. Up until perhaps 30 years ago, it was common for such examinations under anesthesia (EUA) to be done without patient knowledge or consent, but the practice came under ethical and legal scrutiny in the 1980s and 1990s.^4-6^ Patients and medical trainees almost uniformly view consent as necessary for such intimate exams, and professional guidelines recognized the need for prior consent.^7-10^ Nonetheless, prior surveys of practices have shown that consent was not uniformly sought from patients, nor is it legally mandated in all U.S. states, and this situation persists today.^11-19^

While patients express strong preferences to be informed and asked for consent, several small studies suggest that most patients will consent when asked. Bibby and colleagues reported that all 69 patients who were interviewed in their Birmingham (UK) hospital expressed the desire to be informed prior to a medical student EUA.^4^ Wainberg and colleagues surveyed 102 patients at their Calgary clinic, asking about their preferences for being informed and willingness to permit a medical student EUA, reporting that a majority (72%) would want to be asked for consent. Of 93 patients who responded to a question about whether they would consent, 62% indicated they would give permission, 5% indicated they would only permit a female student to perform the EUA, 14% would refuse, and 18% were undecided.^20^ Finally, one secondary source Consent for pelvic exams under anesthesia refers to an unpublished 2021 retrospective chart review in a Chicago family planning clinic that reported 10.4% of patients refused medical student EUAs.^17^

The Department of Obstetrics and Gynecology (ObGyn) in the Perelman School of Medicine at the University of Pennsylvania Health System instituted express consent for medical student EUAs 2 decades ago.21 This was implemented as an electronic consent in 2016. For patients undergoing planned obstetric or gynecological procedures, the surgeon (who may be an attending, fellow, or resident) asks patients if they are willing to allow a medical student to perform an EUA, and the patients’ choice is recorded by capture of text and stored in EPIC back-end tables. While editable, two alternative forms of the last sentence of the text are provided for the physician from which to choose. See Figure 1. It is the purpose of this study to examine patients’ actual choices and perform an exploratory analysis of demographic characteristics of patients and providers and consent to medical student EUAs.

**Figure 1.** Alternative consent and refusal language of the electronic EUA consent form.

## Methods

We performed a single academic institution, multi-site, retrospective chart review, from a large urban teaching hospital. We drew a census from the electronic medical record of EUA consent forms for all patients from whom consent was sought; we planned to search back 4 years to capture data before the Covid-19 pandemic, as concerns about privacy or limiting exposure to unnecessary medical personnel may have been heightened during the pandemic. Consent forms were matched by date and patient Medical Record Numbers to patient demographics (age, race, religion, and insurance carrier) and pre- and post-procedure diagnosis codes, which further enabled linking of physician name. We captured the provider whose EPIC account was open as well as the provider with whom each patient had a scheduled visit at the time EUA consent was sought, providers’ department and specialty, and site location. Records were anonymized by the Penn Medicine Data and Analytics core, and provided to the researchers in anonymous form. Because some text captured patient names, the study was approved by the Penn Institutional Review Board. All identifying information was irreversibly stripped from the records after coding.

Coding of consent form text was initially conducted by the Data and Analytics analyst as part of the data capture; one author manually checked 100% of the census by running text searches for key phrases unique to consent and refusal, as well as related key terms (e.g., male, refus!, decline, not, permit), reading the consent or refusal sentence for every form, and assigning numeric codes to categorical data. Coding from the manual step was checked by a second author.

Finally, to gain some insight into the patients who are asked for the EUA consent, Data and Analytics provided the number of gynecology visits and surgical cases by patient race for the same time period as our data on consent. We chose this sample of cases because these are the patients most likely to be asked for EUA consent.

We planned exploratory assessment using contingency tables and logistic regression to examine potential relationships between patient and provider characteristics and patients’ choices to allow or restrict medical student EUA. All analyses were performed using Stata 12.1 (StataCorp, © 2014). Given the exploratory and hypothesis-generating nature of this analysis, we did not plan to adjust p-values for multiple comparisons.^22^

## Results

Data was captured and downloaded on February 2 and July 31, 2023. While we attempted to search back to 2018 to enable inclusion of patients treated prior to the Covid-19 pandemic, our search method only provided access to consent forms on and after September 20, 2020; this was attributed to a change in system design. We thus captured every use of the EUA consent from September 20, 2020, through December 31, 2022. We have records for 4000 unique patients.

Patients were an average of 45.7 years of age (range: 16-88). There were 16 teenagers (including 5 minors), and 39 patients in their 80s. Patients self-identified predominantly as white (1921, 48.0%), Black or African American (1613, 40.3%), Asian (165, 4.1%), unknown or patient declined (168, 4.2%), and other (including American Indian or Alaskan Native, East Indian, Hispanic/Latino Black, Hispanic/Latino White, and Native Hawaiian or other Pacific Island) (133, 3.3%). Patients also self-identified with 2 dozen religions; grouping faiths showed that just over half were Christian (2003, 50.1%), while religion is undocumented for many patients (1278, 32.0%). Other patients identified as Muslim/Moslem/Islamic (179, 4.5%), Jewish (151, 3.8%), and nondenominational and other (389, 9.7%). Information about insurance coverage or other payors was available for 3331 patients (83.3%); of whom almost two-thirds of patients had commercial health insurance (2170, 65.1%), while the remainder had Medicare, Medicaid, or other public coverage (1161, 34.8%).

Of the 4000 patients, 142 (3.6%) patients were removed from further analysis because of uncertainty about whether they were asked for (and agreed to) EUA consent. For reasons unknown, only truncated consent forms were included in the medical record which did not include the consent or refusal sentences (that is, the italicized sentences in Figure 1 were missing from the EPIC record). Of the remaining 3858 patients, 308 (8.0%) were asked for EUA consent more than once (on different days). Of the 308 patients, 285 were asked twice, 20 were asked 3 times, and 3 were asked 4 times.

Providers were identified in 3798 (98.4%) of the cases in which EUA consent was sought. We identified the provider primarily from the EPIC record identifying the provider who was logged in at the time EUA consent was sought, or, if that person was an ObGyn fellow or resident or a non-ObGyn physician, we identified the ObGyn provider with whom the patient had a scheduled visit at the time EUA consent was sought. This approach enabled us to identify attending physicians for nearly all patients (3610, 95.0%), while residents (152, 3.9%) and fellows (30, 0.8%) made up the remainder. Overall, 140 providers were identified, 23 males and 117 females. Of the 23 males, 10 (43.5%) hold appointments in departments other than ObGyn, while only 8 of 117 female physicians (6.8%) are in other departments (χ2=23.03 w/1 degree of freedom, p<0.001). As this implies, 109 of 122 ObGyn physicians (89.3%) are female. The 18 non-ObGyn physicians were involved in the care of only 26 patients, with 16 of the 18 involved in a single case. Because EUA consent was sought from these patients, we have left them in the census, but because of anonymization of records, we have no ready means for checking why EUA consent was sought in these cases.

The unit of analysis for this study is the patient. We created 2 outcome variables: Consent and Flip. Consent is when a patient allows EUA every time asked. We assess nonconsent as any situation where a patient refuses EUA consent or limits consent to female medical students or otherwise expresses a wish that medical students not take part in their care. Flip is coded only for patients who were asked for EUA consent more than once, and reflects inconsistent choices for patients who sometimes said yes and at other times said no (all of whom were coded as refusers).

Overall, 550 of 3858 patients (14.2%) refused or limited EUA consent. Of the 550, 9 (1.6%) limited consent to only female medical students, including one who gave specific consent for one named female student, and 2 patients (0.4%) refused to have a medical student present at all during their surgery. Of the 308 patients asked more than once, 241 (78.2%) consistently said yes, 21 (6.8%) consistently refused, and 46 (14.9%) were inconsistent. The more times a patient was asked, the more likely she was to refuse EUA consent (Cuzick extension of Wilcoxon rank-sum test for ordered groups, z=2.63, p=0.009).

We performed exploratory univariate and multivariate contingency table and logistic regression analyses to examine whether any of the descriptive factors about patients or their physicians appear related to whether patients consented, and whether those who were asked for EUA consent more than once were consistent. Univariate analysis was highly consistent with multivariate analyses, so here we only present the results of a multiple logistic regression. This yields more robust estimates than univariate analysis, accounting for correlations amongst independent variables. As noted above, given the exploratory nature of this analysis, p-values have not been adjusted for multiple comparisons. Table 1 presents the results of this analysis of Consent. Note that there were no differences across provider department affiliation or specialties, so these variables have been omitted from the model. A similar analysis of Flip found no relationship with any independent variables at lower than a 0.05 level of significance.

**Table 1.** Multiple logistic regression results showing factors influencing the likelihood of pelvic EUA Consent.

| **FACTOR** | **N** | **Consent**<br>**Rate (%)** | **Odds Ratio** | **p-value** |
| --- | --- | --- | --- | --- |
| **Patient Characteristics:** |  |  |  |  |
| Age (in years) | 3858 | – | 1.0036 | 0.332 |
| **Race:** |  |  |  |  |
| White (comparison group) | 1865 | 90.5 | – | – |
| Black or African American | 1541 | 81.1 | 0.482 | <0.001 |
| Asian | 158 | 74.7 | 0.303 | <0.001 |
| Unknown* | 140 | 85.4 | 0.707 | 0.210 |
| Other† | 115 | 86.9 | 0.649 | 0.071 |
| **Religion:** |  |  |  |  |
| Christian (comparison group) | 1935 | 87.1 | – | – |
| Unknown¶ | 1236 | 85.2 | 0.872 | 0.224 |
| Other** | 374 | 86.6 | 1.122 | 0.505 |
| Muslim/Moslem/Islamic | 169 | 72.2 | 0.598 | 0.008 |
| Jewish | 144 | 86.1 | 0.617 | 0.063 |
| **Payors:** |  |  |  |  |
| Commercial (comparison group) | 2113 | 87.1 | – | – |
| Public payors | 1120 | 82.0 | 0.744 | 0.007 |
| Unknown | 625 | 87.8 | 1.026 | 0.858 |
| **Provider Characteristics:** |  |  |  |  |
| **Physician gender:** |  |  |  |  |
| Female (comparison group) | 3262 | 85.2 | – | – |
| Male | 536 | 88.6 | 1.427 | 0.016 |
| **Time of surgery** |  |  |  |  |
| September 2020 – December 2021 | 1841 | 87.4 | – | – |
| January 2022 – December 2022 | 2017 | 84.3 | 0.789 | 0.013 |

Notes: \* Includes Patient Declined.
† Includes American Indian or Alaskan, East Indian, Hispanic Latino, Native Hawaiian or Other Pacific Islander, and Some Other Race.
¶ Includes No Affiliation Given.
\*\* Includes Agnostic, Atheist, Buddhist, and Hindu.

Finally, we were provided raw counts of all patients receiving gynecology care in surgical and nonsurgical visits at least once from September 2020 through December 2022, by patient race. Of the 4000 patients from whom EUA consent was sought, 3527 (88.2%) were drawn from the population of patients identified in EPIC as gynecology patients (i.e., coded for gynecology visits and gynecology surgery, which includes obstetrics). This allows us to estimate the probability of gynecology patients being asked for EUA consent as well as the odds of being asked by racial group. See Table 2.

**Table 2.** The probability (Pr) and numbers of patients asked for EUA consent during gynecology visits and surgery from September 2020 through December 2022, by patient race.

| **Race** | **Gynecology Cases**<br>**N (%)** | **EUA Consent requests**<br>**N (%)** | **Pr (ask for EUA consent) %** | **OR\*** | **p-value** |
| --- | --- | --- | --- | --- | --- |
| White | 20872 (44.9) | 1712 (48.5) | 8.2 | - | - |
| Black or African American | 18869 (40.6) | 1450 (41.1) | 7.7 | 0.932 | <0.057 |
| Asian | 3111 (6.7) | 154 (4.4) | 5.0 | 0.583 | <0.001 |
| Some Other Race | 2057 (4.4) | 86 (2.4) | 4.2 | 0.488 | <0.001 |
| Unknown | 1117 (2.4) | 84 (2.4) | 7.5 | 0.910 | 0.417 |
| All others<sup>†</sup> | 476 (1.0) | 41 (1.2) | 8.6 | 1.054 | 0.747 |
| Totals | 46502 | 3527 | 7.6 |  |  |

Notes: \* Odds of being asked for EUA consent in comparison to White patients.
<sup>†</sup> Includes Patient Declined, Native Hawaiian or Other Pacific Islander, American Indian or Alaskan Native, Hispanic Latino, and East Indian.

## Discussion

Our findings show that overall about 1 in 7 patients refuse consent for medical students to conduct an EUA, generally consistent with other reports.^17,20^ As shown in Table 1, consent rates vary substantially among identified racial groups, ranging from 90.5% of white patients to 81.1% of Black or African American patients and 74.7% of Asian patients. Overall, non-White patients were almost twice as likely as White patients to refuse (18.7% vs. 9.5%). Religious affiliation was also related, with 72.2% of patients identifying as Muslim/Moslem/Islamic consenting. A majority of patients in our sample who identify as Muslim are Black (147 of 169), and a test of the model with this interaction showed that the race and religion main effects remain significant while the interaction is not (p=0.057). Rates of consent amongst patients with other less-prevalent religious affiliations varied from 100.0% of 58 Agnostic or Atheist patients, 71.4% of 21 Hindu patients, to 25% of 4 Christian Scientist patients. As shown in Table 1, patients on Medicare or Medicaid programs were also somewhat less likely to consent, and the rate of consent appears to have dropped in 2022 compared to the earlier period (which was the height of the pandemic).

While patient age was not a significant predictor in our analysis, younger patients were the least likely to agree to medical student EUAs. The highest refusal rate was in patients under 30, with 17.1% of 422 patients saying “no”. This included refusal by 6 of 16 teenagers (37.5%).

Physician gender appears to play a role as well, with male physicians having a slightly lower refusal rate (11.4%) than their female colleagues (14.8%; χχ2=4.48 w/1 degree of freedom, p=0.034). As noted above, the overwhelming majority (89.3%) of ObGyn physicians are female.

We are unable to shed light on the reasons for the observed differences in rates of consent. Patients may distrust their providers, be concerned about their privacy, or simply not want unnamed medical students (of either gender) performing intimate examinations while they are unconscious. Patients may not fully grasp the role medical students may have during surgical procedures, which may involve genital exposure and contact, albeit for therapeutic and not training purposes. Moreover, the finding that male physicians are slightly more likely to secure consent for a medical student EUA raises questions about the consent processes and about the perceived gender and power dynamics in the clinical encounter. It is possible that this finding reflects a selection bias, with patients who choose or accept a referral to a male physician perhaps more likely to agree to a pelvic examination by an unknown, and possibly male, medical student.

Our data on the racial distribution of gynecology patients suggests that patients identifying as Asian or Some Other Race in the medical record were less likely than White patients to be asked for EUA consent. There is no formal process for selection of patients, and we have little insight into why there may be differences in patient selection.

What is the moral significance of higher rates of refusal amongst patients in racial and religious groups? It is broadly (but not yet universally) viewed that consent is ethically necessary, and the fact that some patients would be more likely than others to say no heightens the dignitary harm from a nonconsensual invasion of intimate bodily integrity. The fact that this harm is more likely to fall on persons of color or religious minorities perpetuates the historic wrongs visited upon the vulnerable.^23^ From our perspective, this bolsters the arguments for broader adoption of laws requiring informed consent from patients for nontherapeutic, educational interactions with medical trainees.^14-17^

One primary limitation on this analysis is that the physicians identified in the medical record may not have been the physician seeking EUA consent, and there’s no reliable way to determine from the medical record who was present and sought consent in each case. Nonetheless, the difference shown in rates of consent based on physician gender suggests that some aspect of the clinical encounter influences patients’ choices which could be the focus of future qualitative study. A second limitation is that physicians were not asked their preferred gender identities, with gender coded from each provider’s institutional web page; we believe any imputed error is likely to be small, as a recent study showed that less than 2% of adults (including 5% of young adults under 30) identify as transgender or nonbinary.^24^ A third limitation is that, because we only look at how electronic EUA consents are completed, we know little about the processes followed by physicians in securing the consent and how knowledgeable patients are about their choices. Fourth, while we collected pre- and post-procedure diagnosis codes, we did not attempt to assess whether there were any associations of diagnoses or procedures with rates of consent. We note that the rate of consent was no different between ObGyn and oncology specialists. Fifth, only a portion of patients (88.2%) asked for EUA consent were seen for gynecology visits or surgeries, so we are unsure whether the sample we analyzed is representative of the entire population from which the EUA consent population was drawn. Our analysis also does not account for multiple requests, though patients asked multiple times comprise a small fraction (8.0%) of cases. Finally, our census is limited to patients treated during the Covid-19 pandemic and its aftermath, and our finding that the rate of consent appears to have dropped in 2022 suggests that the pandemic may have had an impact on patients’ willingness to accede to their physicians’ requests, though further research is needed to see if the observed change persists.

In conclusion, we find that a majority of patients will permit medical students to conduct a pelvic EUA, when asked for their consent, but that some patients are substantially more likely to refuse than others. Patient’s rights to control over their own bodies can only be respected if their physicians take seriously the ethical obligation to inform their patients and ask them for permission.

INFORMATION AND CONSENT FOR PELVIC EXAMINATION UNDER GENERAL ANESTHESIA After you are put to sleep with general anesthesia, A PELVIC EXAM may be performed as part of your planned gynecologic surgery. This examination is a standard part of many gynecologic procedures. Often, it provides the surgeon with valuable information for the safe conduct of the surgery. For example, findings on a pelvic examination might be used to determine the best surgical approach for your particular problem or the optimal location of the incision for your surgery. If a pelvic examination is planned as part of your surgical procedure, your physician will include this in the description of the surgical procedure for which you provide written consent. If a pelvic examination is performed after you are under general anesthesia, you will be draped in a manner similar to an examination in the office. Your surgeon may ask another physician member of the team to also perform a pelvic examination to confirm the findings or to provide an additional opinion. Performing a pelvic examination under anesthesia may also provide your surgeon with an opportunity to teach his/her surgical assistants regarding how to properly perform the procedure. If the surgical team will include a medical student, your physician also may ask you to read this information sheet and decide if you will permit a medical student, who will be assisting with the surgical procedure, to also perform a pelvic examination. Under no circumstances, will a medical student perform a pelvic examination under anesthesia without your permission. If you decide that you do not want a medical student to perform this examination, this decision will not affect your care in any way. I have read and understand this information sheet and my questions have been answered to my satisfaction. I certify that my physician has informed me that a pelvic examination is planned or may be necessary as part of my surgical procedure and I consented to this examination.*

## Data Availability

Anonymized data analyzed for this study are available upon reasonable request to the authors.

## Acknowledgements

The authors thank Penn Data and Analytics and Ting-shan Chiu for their assistance in assembling the information used in this study, and Kavita Shah Arora, Andrew Fesnak, and several anonymous reviewers for comments. Responsibility for the study is solely that of the authors.

There was no funding for this study.

## Consent

*I understand that a medical student may assist my physician with the surgery. I hereby consent to permit a medical student who is part of the surgical team for my procedure to perform a pelvic examination under anesthesia as part of my procedure under the direct supervision of my physician*.

## Refuse

*If a medical student is part of the surgical team, I request that the student not be permitted to perform a pelvic exam as part of my procedure*.

### Notes

* The alternative italicized sentences appear without break in the paragraph, in normal (not italicized) font. This text is fully editable by the surgeon at the time consent is discussed with each patient.

